# On the frontlines in Shanghai: Stress, burnout and perceived benefit among COVID-19 testers and other personnel during the Omicron wave lockdown

**DOI:** 10.1101/2022.10.25.22281504

**Authors:** Zhimin Xu, Xia Liu, Gabriela Lima de Melo Ghisi, Lixian Cui, Sherry L Grace

## Abstract

**Background:** COVID-19 control measure stringency including testing has been among the highest globally in China. Psychosocial impact on pandemic workers in Shanghai, and their pandemic-related attitudes were investigated.

**Methods:** Participants in this cross-sectional study were healthcare providers (HCP) and other support workers. A Mandarin self-report survey was administered via Wenjuanxing between April-June 2022 during the omicron-wave lockdown. The Perceived Stress Scale (PSS) and Maslach Burnout Inventory were administered, as well as pandemic-specific questions.

**Results:** 887 workers participated, of which 691 (77.9%) were HCPs. They were working a mean of 6.25±1.24 days/week for 9.77±4.28 hours/day. Most participants were burnt-out, with 143(16.1%) moderately and 98(11.0%) seriously. Total PSS was 26.85±9.92/56, with 353(39.8%) participants having elevated stress. Workers perceived their families primarily as fully supportive (n=610, 68.8%), or also extremely concerned (*n*=203, 22.9%). Most wanted counselling and stress relief, but half(*n*=430) reported no time for it; indeed, 2/3rds wanted a few days off to rest (*n*=601).Many workers perceived benefits: that they fostered more cohesive relationships (*n*=581, 65.5%), they will be more resilient (*n*=693, 78.1%), and were honored to serve (*n*=747, 84.2%).Negative impacts were greater in HCPs, those with economic insecurity, and that did not perceive benefit (all *p*<.05).In adjusted analyses, those perceiving benefits showed significantly less burnout (OR=0.573, 95% CI=0.411 - 0.799), among other correlates.

**Conclusions:** Pandemic work, including among non-HCP, is stressful, but some can derive benefits.

## INTRODUCTION

The COronaVIrus Disease (COVID-19) pandemic has resulted in major negative impacts for economies, health systems and citizens worldwide.^1^ While the impact on the health of all people has been of major concern, particularly before the availability of vaccines or any treatment, it has been a particular concern for those working on the front lines.^2^ While most who work in caring professions derive great satisfaction from their work, health care providers (HCPs) and other front-line personnel (e.g., emergency services, testing) are at risk of infection while performing their duties (and hence infecting loved ones when they return home), workloads and uncertainty have increased exponentially, and mental health has suffered.^3,4^

Measures to control infection spread include testing. In China, universal mass testing is undertaken in accordance with the “dynamic zero-COVID” policy.^5–7^ Along with the strict lockdowns, this likely represents amongst the greatest stringency in COVID-19 control measures of any country globally.^8^ All who test positive must isolate at central quarantine centers or hospitals; many mobile centers have been rapidly built. Moreover, “anti-epidemic packages” are distributed broadly, including test kits, masks, and herbal pills.^6^ Little is known about the perspectives and well-being of these workers after all these measures were implemented.

The negative economic impact of the control policies has been great globally, but also in China given the stringency. Moreover, in locked-down compounds, people are also struggling to secure what have become scarce food and medical resources and getting medical attention for any cause is challenging; population acceptance of and reaction to the imposed control measures has been fraught. Residents have expressed their frustration to pandemic workers;^7^ for example, at one temporary quarantine center in Shanghai, “a video circulating on Chinese social media showed an angry crowd confronting a hazmat-suited worker”.^6^ It was reported that over half of a construction crew building a temporary hospital contracted COVID-19, angering local residents, highlighting the health risks to pandemic workers and the negative context in which they are working. Other video footage has circulated of “people yelling in frustration from their balconies”.^5^

A recent article in the *Journal of the American Medical Association* calls for more attention to well-being not just in physicians and nurses, but throughout the healthcare workforce, and the urgent need to measure this.^2^ Therefore, the objective of this study was to assess the well-being and perceptions of those involved in pandemic testing during the most recent Omicron wave in Shanghai, including their stress and burnout, but also any perceived benefits of their work. The associations between these with degree of economic security, type of worker (HCP vs other), and perception of benefit were also tested.

## METHODS

### Design and Procedure

The study was approved by the Xinhua Hospital research ethics board (XHEC-C-2022-042). The anonymous online survey was created by the Psychosomatic Medicine Committee of the Shanghai Association of Integrated Traditional Chinese and Western Medicine in Mandarin. The survey was prefaced by consent.

Data collection for this cross-sectional study was undertaken between April-June 2022 via the online survey platform Wenjuanxing (https://www.wjx.cn/). The questionnaire was distributed by the Community Doctor Group of the Psychosomatic Medicine Special Committee of the Shanghai Society of Integrated Chinese and Western Medicine. The doctors distributed the questionnaire to their hospitals’ anti-COVID WeChat groups and their communities’ anti-COVID worker WeChat groups.

### Setting and Participants

Shanghai is a cosmopolitan city of 25 million people. The city was in the midst of localized restrictions from March 2022 due to the highly transmissible omicron variant and was in full lockdown from April 5 for two weeks. Each day residents were required to do COVID-19 nucleic acid testing. HCP including physicians and nurses were tasked with testing building by building, and other support staff or volunteers (e.g., full-time community workers, political party staff, police officers, community residents) helped with scanning bar codes, disinfection, distribution of anti-epidemic materials, as well as fetching medicine for residents, etcetera.

Participant inclusion criteria were HCP doing screening or involved in direct COVID-19 patient care in the mobile hospitals or other support staff or volunteers participating in activities as outlined above, in communities with strict lockdown. Participant exclusion criteria were HCP not working in the frontline against COVID-19; HCP having history of mental illness.

### Measures

All items were self-reported. Non-psychometrically-validated items were generated by the Psychosomatic Medicine committee. For instance, the three perceived benefit items queried about better relationships, greater resilience, and honor in serving, with yes/no response options.

-

The Maslach Burnout Inventory General Survey (MBI-GS) was designed for use with occupational groups other than human services and education;^9^ the psychometrically validated Chinese version was used in this study.^10,11^ This 15 item questionnaire consists of three subscales: emotional exhaustion (5 items), cynicism (4 items), and professional efficacy (6 items). Respondents rated how often they experienced each from a list of symptoms on a 7-point scale ranging from 0=never to 6=every day. The degree of burnout is determined to be higher when the scores in exhaustion and cynicism are high, and the score for professional efficacy is low.^9^ For the Chinese version, it is not suggested to use total score, but scores the top one-third of each dimension were used as a cutoff for burnout (in this data, emotional exhaustion 20 points, dehumanization 13 points, personal achievement 18 points). Moreover, burnout was categorized as: no burnout (scores on three dimensions are lower than the cut-off value), slight burnout (scores on one dimension are higher than the cut-off value), moderate burnout (the scores on two dimensions are higher than the cut-off value) and serious burnout (the scores on three dimensions are higher than the cut-off value).

Psychological stress was measured using the Chinese version of the Perceived Stress Scale (CPSS), which has been psychometrically validated with good validity and reliability demonstrated.^12,13^ The 14-item self-reported questionnaire was designed to measure the degree to which situations in one’s life are appraised as stressful. Respondents answer the 14 questions using a 5-point Likert-type scale ranging from 0=never and 4=very often. Two subscales are evaluated: perceived stress and lack of control. CPSS scores range from 0 to 56; total scores of 0-28 indicate normal levels of stress, 29-42 indicates high stress, and 43-56 indicates excessive stress.

### Statistical Analyses

Descriptive analysis was performed using IBM SPSS statistics v25. The association between attitudes, stress and burnout with type of pandemic worker, perceived economic security and perception of honour in pandemic work were tested using *t*-tests or chi-square analyses, as applicable. Finally, significant correlates of the latter were tested using logistic regression analysis.

## RESULTS

Of 887 responding participants, most were female, married HCPs (Table 1). There were also volunteers, and other support workers such as local police and members of the political party. They were working on average more than six days/week for almost ten hours/day. More than half reported being negatively economically impacted by the pandemic, and half were concerned about their future working prospects. More than half were worried about being infected, but worked anyways (Table 2).

**Table 1:**
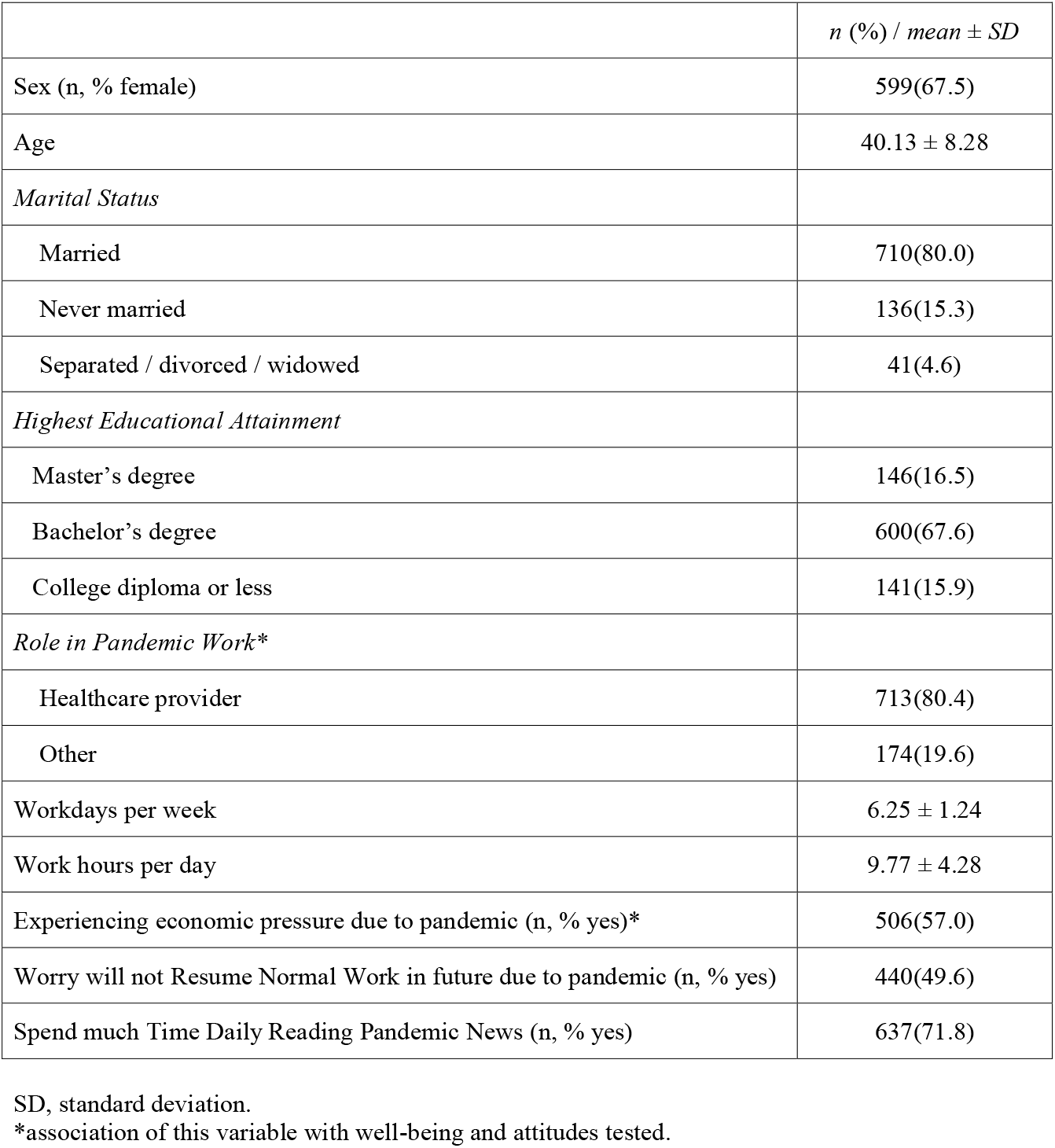
Participant Sociodemographic and Occupational Characteristics, *N*=887

|  | *n (%) / mean ± SD* |
| --- | --- |
| Sex (n, % female) | 599(67.5) |
| Age | 40.13 ± 8.28 |
| *Marital Status* |  |
| Married | 710(80.0) |
| Never married | 136(15.3) |
| Separated / divorced / widowed | 41(4.6) |
| *Highest Educational Attainment* |  |
| Master's degree | 146(16.5) |
| Bachelor's degree | 600(67.6) |
| College diploma or less | 141(15.9) |
| *Role in Pandemic Work\** |  |
| Healthcare provider | 713(80.4) |
| Other | 174(19.6) |
| Workdays per week | 6.25 ± 1.24 |
| Work hours per day | 9.77 ± 4.28 |
| Experiencing economic pressure due to pandemic (n, % yes)* | 506(57.0) |
| Worry will not Resume Normal Work in future due to pandemic (n, % yes) | 440(49.6) |
| Spend much Time Daily Reading Pandemic News (n, % yes) | 637(71.8) |

SD, standard deviation.
\*association of this variable with well-being and attitudes tested.

**Table 2:** Participant Stress, Burnout and Attitudes related to the Pandemic, *N*=887

|  | *n* (%) /<br>mean ±<br>*SD* | Association with Type of Worker |  |  |  | Association with Economic Security |  |  |  | Association with Perceived Benefit / Honor |  |  |  |
| --- | --- | --- | --- | --- | --- | --- | --- | --- | --- | --- | --- | --- | --- |
|  |  | HCP<br>(*n*=713) | Other<br>(*n*=174) | χ<sup>2</sup>/t | *p* | Not<br>(*n*=506) | Secure<br>(*n*=381) | χ<sup>2</sup>/t | *p* | Yes<br>(*n*=747) | No<br>(*n*=140) | χ<sup>2</sup>/t | *p* |
| *Perception of Familial Attitude Regarding their Pandemic Work* |  |  |  |  |  |  |  |  |  |  |  |  |  |
| Full support | 610<br>(68.8) | 492<br>(69.0) | 118 (67.8) | 6.706 | 0.092 | 311<br>(61.5) | 299<br>(78.5) | 31.833 | **<0.001** | 549<br>(73.5) | 61 (43.6) | 63.593 | **<0.001** |
| Supportive, but<br>extremely<br>concerned | 203<br>(22.9) | 169<br>(23.7) | 34 (19.5) |  |  | 142<br>(28.1) | 61 (16.0) |  |  | 159<br>(21.3) | 44 (31.4) |  |  |
| Neutral | 68 (7.7) | 48 (6.7) | 20 (11.5) |  |  | 47<br>(9.3) | 21 (5.5) |  |  | 36 (4.8) | 32 (22.9) |  |  |
| Full objection | 6 (0.7) | 4 (0.6) | 2 (1.1) |  |  | 6 (1.2) | 0 |  |  | 3 (0.4) | 3 (2.1) |  |  |
| *Perception of Neighbors Attitude Regarding their Pandemic Work* |  |  |  |  |  |  |  |  |  |  |  |  |  |
| Avoidance and<br>discrimination | 91<br>(10.3) | 81<br>(11.4) | 10 (5.7) | 22.247 | **<0.001** | 76 (15.0) | 15 (3.9) | 36.396 | **<0.001** | 65 (8.7) | 26 (18.6) | 40.722 | **<0.001** |
| Same as usual | 424<br>(47.8) | 360<br>(50.5) | 64 (36.8) |  |  | 248<br>(49.0) | 176<br>(46.2) |  |  | 336<br>(45.0) | 88 (62.9) |  |  |
| Encouraging and<br>supportive | 372<br>(41.9) | 272<br>(38.1) | 100 (57.5) |  |  | 182<br>(36.0) | 190<br>(49.9) |  |  | 346<br>(46.3) | 26 (18.6) |  |  |
| *Perceived Stress* |  |  |  |  |  |  |  |  |  |  |  |  |  |
| Normal | 534<br>(60.2) | 407<br>(57.1) | 127 (73.0) | 14.788 | 0.001 | 245<br>(48.4) | 289<br>(75.9) | 72.616 | **<0.001** | 503<br>(67.3) | 31 (22.1) | 156.045 | **<0.001** |
| High pressure | 290<br>(32.7) | 251<br>(35.2) | 39 (22.4) |  |  | 207<br>(40.9) | 83 (21.8) |  |  | 220<br>(29.5) | 70 (50.0) |  |  |
| Excessive pressure | 63 (7.1) | 55 (7.7) | 8 (4.6) |  |  | 54 (10.7) | 9 (2.4) |  |  | 24 (3.2) | 39 (27.9) |  |  |
| *Burnout* |  |  |  |  |  |  |  |  |  |  |  |  |  |
| No burnout | 463<br>(52.2) | 356<br>(49.9) | 107 (61.5) | 12.007 | **0.007** | 209<br>(41.3) | 254<br>(66.7) | 66.352 | **<0.001** | 438<br>(58.6) | 25 (17.9) | 224.226 | **<0.001** |
| Slight burnout | 183<br>(20.6) | 146<br>(20.5) | 37 (21.3) |  |  | 112<br>(22.1) | 71 (18.6) |  |  | 168<br>(22.5) | 15 (10.7) |  |  |
| Moderate burnout | 143<br>(16.1) | 123<br>(17.3) | 20 (11.5) |  |  | 109<br>(21.5) | 34 (8.9) |  |  | 105<br>(14.1) | 38 (27.1) |  |  |
| Serious burnout | 98<br>(11.0) | 88<br>(12.3) | 10 (5.7) |  |  | 76 (15.0) | 22 (5.8) |  |  | 36 (4.8) | 62 (44.3) |  |  |
| Critical; need<br>consultation<br>(>100) | 37 (4.2) | 34 (4.8) | 3(1.7) |  |  | 32(6.3) | 5(1.3) |  |  | 7 (0.9) | 30 (21.4) |  |  |
| *Perceive life will*<br>*return to normal*<br>*post-pandemic (n,*<br>*% yes)* | 691<br>(77.9) | 551<br>(77.3) | 140 (80.5) | 0.822 | 0.415 | 358<br>(70.8) | 333<br>(87.4) | 35.005 | **<0.001** | 627<br>(83.9) | 64 (45.7) | 100.058 | **<0.001** |
| *Perceived Benefits from Pandemic Work (n, % yes)* |  |  |  |  |  |  |  |  |  |  |  |  |  |
| More cohesive<br>community<br>relationships | 581<br>(65.5) | 442<br>(62.0) | 139 (79.9) | 19.818 | **<0.001** | 292<br>(57.7) | 289<br>(75.9) | 31.669 | **<0.001** | 556<br>(74.4) | 25 (17.9) | 166.996 | **<0.001** |
| I will be more<br>resilient in future | 693<br>(78.1) | 537<br>(75.3) | 156 (89.7) | 16.831 | **<0.001** | 354<br>(70.0) | 339<br>(89.0) | 45.994 | **<0.001** | 652<br>(87.3) | 41 (29.3) | 232.084 | **<0.001** |
| Feel honored to | 747 | 590 | 157 (90.2) | 5.889 | **0.015** | 403 | 344 | 18.527 | **<0.001** |  |  |  |  |
| participate in fight against COVID-19* | (84.2) | (82.7) |  |  |  | (79.6) | (90.3) |  |  |  |  |  |  |
| *Regret working in the fight against COVID-19* (n, % yes) | 48 (5.4) | 39 (5.5) | 9 (5.2) | 0.024 | 0.876 | 39 (7.7) | 9 (2.4) | 12.132 | **<0.001** | 6 (0.8) | 42 (30.0) | 196.354 | **<0.001** |
| *Committed to continue pandemic work* (n, % yes) | 772 (87.0) | 616 (86.4) | 156 (89.7) | 1.317 | 0.251 | 422 (83.4) | 350 (91.9) | 13.800 | **<0.001** | 697 (93.3) | 75 (53.6) | 164.971 | **<0.001** |
| *Concern about Infection at Work* |  |  |  |  |  |  |  |  |  |  |  |  |  |
| Always worried and scared | 116 (13.1) | 99 (13.9) | 17(9.8) | 2.202 | 0.332 | 90 (17.8) | 26 (6.8) | 32.472 | **<0.001** | 76 (10.2) | 40 (28.6) | 39.515 | **<0.001** |
| I am worried, but someone needs to do this job | 500 (56.4) | 400 (56.1) | 100 (57.5) |  |  | 290 (57.3) | 210 (55.1) |  |  | 425 (56.9) | 75 (53.6) |  |  |
| With the risk protection measures and equipment, I do not worry about infection | 271 (30.6) | 214 (30.0) | 57 (32.8) |  |  | 126 (24.9) | 145 (38.1) |  |  | 246 (32.9) | 25 (17.9) |  |  |
| *Biggest Concerns in Pandemic Work* |  |  |  |  |  |  |  |  |  |  |  |  |  |
| Limited ability and energy to complete tasks assigned by superiors | 315 (35.5) | 271 (38.0) | 44 (25.3) | 9.883 | **0.002** | 207 (40.9) | 108 (28.3) | 14.978 | **<0.001** | 237 (31.7) | 78 (55.7) | 29.623 | **<0.001** |
| Doing my best, but the residents do not understand | 410 (46.2) | 332 (46.6) | 78 (44.8) | 0.170 | 0.680 | 269 (53.2) | 141 (37.0) | 22.817 | **<0.001** | 333 (44.6) | 77( 55.0) | 5.152 | **<0.001** |
| So physically fatigued, I cannot continue | 331<br>(37.3) | 285<br>(40.0) | 46 (26.4) | 10.954 | **0.001** | 230<br>(45.5) | 101<br>(26.5) | 33.350 | **<0.001** | 254<br>(34.0) | 77 (55.0) | 22.223 | **<0.001** |
| Leadership direction is unclear, so work is disorganized | 328<br>(37.0) | 264<br>(37.0) | 64 (36.8) | 0.004 | 0.952 | 217<br>(42.9) | 111<br>(29.1) | 17.637 | **<0.001** | 245<br>(32.8) | 83 (59.3) | 35.496 | **<0.001** |
| We do not have the supplies we need to complete our tasks | 294<br>(33.1) | 228<br>(32.0) | 66 (37.9) | 2.237 | 0.135 | 197<br>(38.9) | 97(25.5) | 17.806 | **<0.001** | 228<br>(30.5) | 66 (47.1) | 14.698 | **<0.001** |
| Feel caught between leadership and the people, and cannot please either | 242<br>(27.3) | 191<br>(26.8) | 51 (29.3) | 0.448 | 0.503 | 169<br>(33.4) | 73(19.2) | 22.212 | **<0.001** | 185<br>(24.8) | 57 (40.7) | 15.116 | **<0.001** |
| Some people will not cooperate with us, raising difficulties | 471<br>(53.1) | 381<br>(53.4) | 90 (51.7) | 0.165 | 0.685 | 286<br>(56.5) | 185<br>(48.6) | 5.537 | **0.019** | 396<br>(53.0) | 75 (53.6) | 0.015 | 0.903 |
| Other (no concern) | 99<br>(11.2) | 75<br>(10.5) | 24 (13.8) | 1.512 | 0.219 | 55 (10.9) | 44 (11.5) | 0.101 | 0.751 | 88 (11.8) | 11 (7.9) | 1.830 | 0.176 |
| *Want Psychological Counselling and/or Stress Management Supports* |  |  |  |  |  |  |  |  |  |  |  |  |  |
| Yes | 197<br>(22.2) | 150<br>(21.0) | 47(27.0) | 2.890 | 0.236 | 108<br>(21.3) | 89 (23.4) | 12.670 | **0.002** | 178<br>(23.8) | 19 (13.6) | 9.726 | **0.008** |
| Willing, but have no time | 430<br>(48.5) | 351<br>(49.2) | 79(45.4) |  |  | 270<br>(53.4) | 160<br>(42.0) |  |  | 362<br>(48.5) | 68 (48.6) |  |  |
| No | 260<br>(29.3) | 212<br>(29.7) | 48(27.6) |  |  | 128<br>(25.3) | 132<br>(34.6) |  |  | 207<br>(27.7) | 53 (37.9) |  |  |

| *Current Need to Support Your Pandemic Work* |  |  |  |  |  |  |  |  |  |  |  |  |  |
| --- | --- | --- | --- | --- | --- | --- | --- | --- | --- | --- | --- | --- | --- |
| Few days off to rest | 60<br>(67.8) | 510<br>(71.5) | 91(52.3) | 23.674 | **<0.001** | 353<br>(69.8) | 248<br>(65.1) | 2.171 | 0.141 | 501<br>(67.1) | 100<br>(71.4) | 1.026 | 0.311 |
| Need supplies and better infrastructure for pandemic prevention activities | 372<br>(41.9) | 295<br>(41.4) | 77(44.3) | 0.476 | 0.490 | 242<br>(47.8) | 130<br>(34.1) | 16.766 | **<0.001** | 313 41.9) | 59 (42.1) | 0.003 | 0.958 |
| Need to improve the current quarantine process and strategy | 557<br>(62.8) | 444<br>(62.3) | 113(64.9) | 0.427 | 0.513 | 333<br>(65.8) | 224<br>(58.8) | 4.582 | **0.032** | 458<br>(61.3) | 99 (70.7) | 4.461 | **0.035** |
| Reduced workload | 366<br>(41.3) | 324<br>(45.4) | 42(24.1) | 26.191 | **<0.001** | 244<br>(48.2) | 122<br>(32.0) | 23.536 | **<0.001** | 291<br>(39.0) | 75 (53.6) | 10.392 | **0.001** |
| Financial compensation commensurate with workload | 506<br>(57.0) | 442<br>(62.0) | 64(36.8) | 36.277 | **<0.001** | 342<br>(67.6) | 164<br>(43.0) | 53.435 | **<0.001** | 415<br>(55.6) | 91 (65.0) | 4.292 | **0.038** |

### Impact of Pandemic Work

As shown in Table 2, approximately half of participants were burnt-out, with a substantial proportion severely burnt-out, and some critically so. Mean stress was high. Most workers perceived their families primarily as fully supportive, or supportive and extremely concerned. Over 10% perceived their neighbours were discriminatory and avoiding them, and were very worried about getting infected. Most wanted counselling and stress relief, but half reported no time for it.

As shown in Table 2 as well, their greatest concerns regarding their pandemic work were: lack of cooperation and difficulty raised by some residents, that residents do not understand the work they are doing, lack of physical energy, lack of leadership leading to disorganization, lack of ability and energy to complete assigned tasks, lack of infrastructure, following by feeling like they are caught between leadership and residents and that they cannot please either party; each concern was endorsed by over a quarter of respondents. As also shown in Table 2, what pandemic workers reported they needed most, in descending order of frequency, was: a few days off to rest, improvement in the quarantine process, and financial compensation commensurate with their efforts; each was endorsed by more than half of respondents.

Moreover, economically insecure pandemic workers, who were more often male (*p*=.01) and younger (*p*=.005), perceived significantly less familial and neighbour support, and reported significantly greater stress and burnout (Table 2). They were significantly less likely to perceive life would return to normal, and they perceived less benefit and honor in their work. They had significantly more regret, worry, and endorsed each concern to a significantly greater degree than their economically-secure counterparts. They were significantly more likely to want counselling but not have the time. Finally, they endorsed all needs except for having a few days off than their economically secure counterparts.

### Benefits and Honour in Pandemic Work

Many testers perceived benefit through their work, namely that, they feel honored to serve (*n*=747, 84.2%), through the community fight they will be more resilient in future (*n*=693, 78.1%), and they have fostered more cohesive relationships (*n*=581, 65.5%; Table 2). There was scant regret for pandemic work, and almost 90% said they would continue their work. Almost 80% perceived their life would return to normal post-pandemic.

As shown in Table 2, most of the pandemic attitudes and well-being indicators were also significantly associated with perceiving honour in their pandemic work. In adjusted analyses controlling for sex, age, educational attainment, type of worker and economic pressure, perceiving honor in their pandemic work was significantly associated with age (OR=0.937, 95% CI=0.903-0.972; *p*=0.001), supporting family attitude (OR=1.838, 95% CI=1.208-2.798; *p*=0.004), burnout (OR=0.573, 95% CI=0.411-0.799; *p*=0.001), regret in their work (OR=0.026, 95% CI=0.006-0.112; *p*<.001), as well as were more likely to perceive more cohesive community relationships (OR=4.431, 95% CI=2.277-8.622; *p*<0.001), more resilient in future (OR=5.650, 95% CI=3.014-10.590; *p*<0.001) and concern about physical fatigue (OR=2.093, 95% CI=1.065-4.114; *p*=0.032) (Figure 1).

**Figure 1:**
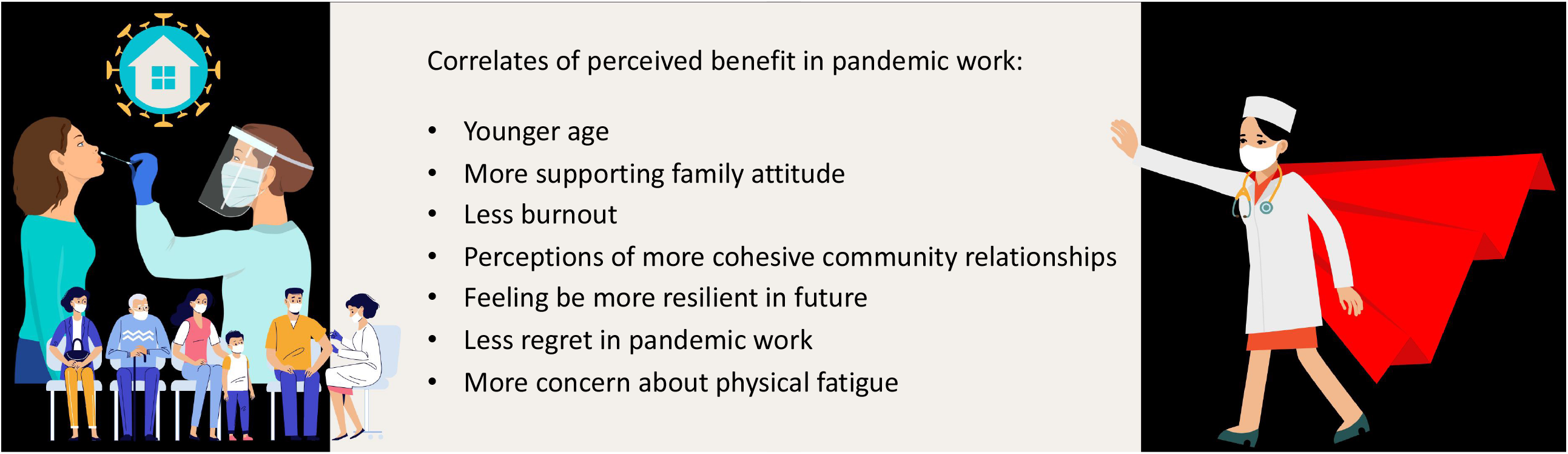
Correlates of Perceived Honor in Pandemic Work Some pandemic workers were more likely to perceive honour in their jobs, and this had several correlates.

### Differences by Type of Pandemic Worker

As also shown in Table 2, HCPs perceived they were more often avoided and less supported by neighbours than other participants, and had significantly higher levels of stress and burnout. They perceived significantly less benefit and honour in their work. They were significantly more concerned about having enough energy to work and their physical fatigue. They were significantly more likely to report they needed a few days off to rest, reduced workload and fairer compensation than non-HCP pandemic workers.

As shown in Table 3, non-HCP pandemic workers were less likely to be female (OR=0.331, 95% CI=0.225-0.487; *p*<0.001), significantly more divorced/widowed (OR=2.329, 95% CI=1.042-5.204; *p*=0.039), showed lower educational attainment (OR=0.582, 95% CI=0.419-0.809; *p*=0.001), fewer working days per week (OR=0.729, 95% CI=0.633-0.840; *p*<0.001) and perceived more supporting attitude from neighbors (OR=1.572, 95% CI=1.134-2.180; *p*=0.007). The overall model was significant (*p*<0.001).

**Table 3:** Multivariate Logistic Regression Analysis of Correlates of Type of Pandemic Worker (*N*=887)

| Independent variable | OR | 95% CI | *p* |
| --- | --- | --- | --- |
| Sex | 0.331 | 0.225—0.487 | <0.001 |
| Age | 0.979 | 0.952—1.007 | 0.144 |
| Marital Status |  |  |  |
| Married | 1 |  |  |
| Never married | 1.130 | 0.612—2.087 | 0.697 |
| Separated / divorced / widowed | 2.329 | 1.042—5.204 | 0.039 |
| Highest Educational Attainment | 0.582 | 0.419—0.809 | 0.001 |
| Economic security | 1.192 | 0.781—1.819 | 0.415 |
| Workdays per week | 0.729 | 0.633—0.840 | <0.001 |
| Hours worked per day | 1.022 | 0.975—1.070 | 0.366 |
| Worry will not Resume Normal Work in future due to pandemic |  |  |  |
| Not Resume Normal Work | 1 |  |  |
| Will Resume Normal Work | 1.029 | 0.667—1.588 | 0.896 |
| Perception of Neighbors Attitude Regarding their Pandemic Work | 1.572 | 1.134—2.180 | 0.007 |
| CPSS levels | 0.957 | 0.617—1.486 | 0.846 |
| MBS levels | 0.991 | 0.751—1.306 | 0.946 |
| Benefit: more cohesive relationships | 1.454 | 0.859—2.462 | 0.163 |
| Benefit: more resilience | 1.689 | 0.840—3.394 | 0.141 |
| Benefit: honor in work | 0.669 | 0.314—1.426 | 0.298 |
| Concern about ability to complete work | 0.720 | 0.459—1.131 | 0.154 |
| Concern about physical fatigue | 0.932 | 0.592—1.467 | 0.762 |
| Constant | 3.804 |  | 0.150 |

OR, odds ratio; CI, confidence interval.
Note: HCP=0, supporting staff=1
Regard CPSS levels and MBS levels as consecutive variables, the higher the score, the more serious the symptoms; Regard highest education attainment and perception of neighbors' attitude as consecutive variables, the higher the score, the higher education or more positive attitude.
Implement "Enter" logistic regression
Omnibus Tests of model Coefficients: $\chi^2=107.601$ , $p<0.001$
-2 log likelihood= 693.999
Cox& Snell $R^2=0.124$ ; Nagelkerke $R^2=0.198$
Hosmer-Lemesho: $\chi^2=2.686$ , $p=0.953$

## DISCUSSION

Given the population of China, surges in COVID-19 cases have the potential to overwhelm healthcare and other systems in the country. While there have been questions about the “zero-COVID” policy, it is a globally-unique approach of mass testing and isolation, that negatively impacts citizens, and requires massive human resources to implement. This is one of the first studies to survey pandemic workers (not just physicians or nurses) in China regarding their perceptions and well-being, and to consider economic impact and perceived benefit. It was undertaken two years into the pandemic at the height of highly-transmissible omicron sub-variant wave. Consistent with findings of HCP in other countries,^14–17^ results show elevated stress and burnout, but also commitment and honor in their pandemic work.

HCP working in the pandemic are significantly more negatively impacted than other workers, which could be due to greater pressure as their work can have life or death impact, yet they have little control over these outcomes. The majority of pandemic workers were economically insecure due to the pandemic itself, so may have been working in such risky and unpopular occupations to better establish their economic security. Nevertheless, the resilience and honor reported – although it could potentially be explained by cognitive dissonance – ^18,19^ was encouraging.

Indeed, this is one of the few studies examining perceived benefit of pandemic work. Many HCP have put their lives and that of their families at risk, and many have died.^20,21^ They have worked under very difficult circumstances, including donning and doffing personal protective equipment, and treating dying patients separated from loved ones. Clearly this takes commitment and dedication, and results of this study demonstrate the honor they feel, the strength they derive from it, including in terms of relationships with others. While early in the pandemic there was much gratitude shown towards HCP, pandemic workers implementing control policies that are disruptive to individuals are often not appreciated, as exemplified in the introduction.

Thus, study implications relate to ensuring workers have what they need to be well, and to indeed derive some satisfaction or meaning in their potentially life-threatening work. What they most wanted was some days off, which is warranted given they are working on average almost 10 hours per day, over 6 days per week. They also desire better quarantine strategies and processes, and compensation commensurate with their workload. With regard to the former, there has been discussion of whether China may re-visit their “zero COVID” policy and what the ramifications would be.^5,22^ With regard to the latter, personal communication from some HCP on the frontline suggested they perceived their salary was low not only considering the context in which they were working, but also because they have to stay in a hotel and they will not get paid for the quarantine. Moreover, government and hospitals promised a bonus, but some have not yet been compensated because hospitals are low on money due to low care volumes related to the pandemic. Other research has identified factors that mediate degree of well-being and burnout of HCP responding to pandemics,^23^ as well as psychological support interventions for them.^24,25^ More research such as this is needed on the psychosocial impact on non-HCP pandemic workers of implementing disliked COVID-19 control policies.

Caution is warranted when interpreting these results. First, this is a cross-sectional study, so the design precludes causal determinations or of direction of effect, and we cannot test how well-being in these workers differs from pre-pandemic levels, early in the pandemic or when there are not active outbreaks and lockdowns. Second, the results may not be generalizable beyond China with its specific political and cultural context and the city of Shanghai more specifically, and where stringency of control measures is great. Moreover, similarity of the sample to the larger population is not known given the recruitment strategy, designed to quickly secure data during the lockdown, but where full reach and non-responders were not codified.

Third, all data were self-report, and hence there may have been socially-desirable responding or other measurement error, and psychological status was not assessed through structured clinical interview. Fourth, multiple comparisons were performed, inflating potential type 1 error.

In conclusion, there has been insufficient attention to the psychosocial well-being of non-HCPs, the impact of waves and associated control measures, as well as the economic stresses and perceived benefits surrounding pandemic work motivation. This study establishes the economic stresses and psychosocial impact among pandemic workers (HCPs and non-alike) are significant during stringent control measures. Continued efforts to improve working conditions and provide evidence-based psychosocial supports are warranted.

## Data Availability

All data produced in the present study are available upon reasonable request to the authors

